# Development and External Validation of a Machine Learning Model for 10-Year Ischemic Stroke Risk Prediction in Diverse Populations

**DOI:** 10.64898/2026.06.22.26356280

**Authors:** Ahmed Khattab, Zhe Wang, Vinodh Srinivasasainagendra, Hemant K. Tiwari, Ruth Loos, Nita Limdi, Marguerite Ryan Irvin

**Author notes:** **<u>Address all correspondence to:</u>** Marguerite Ryan Irvin, Ph.D. 665 University Blvd, Birmingham, AL 35233.

## Abstract

**Importance:** Machine-learning models for ischemic stroke risk prediction are rarely validated across ancestrally distinct cohorts, and the contributions of polygenic risk scores (PRS) and self-reported race in such models remain unclear.

**Objective:** To develop and externally validate a 10-year ischemic stroke risk model and quantify the incremental contributions of laboratory trajectories, PRS, and self-reported race and ethnicity across populations.

**Design, Setting, and Participants:** Retrospective cohort study with model development in the All of Us (AoU) Research Program (n = 34,987; 1,920 incident strokes) and external validation in the Bio*Me* Biobank at Mount Sinai (n = 10,693; 107 incident strokes). Adults aged ≥45 years with ≥1 year of pre-baseline electronic health record data were anchored to a January 2010 baseline with 10-year follow-up.

**Exposures:** Three XGBoost model tiers added laboratory feature trajectories (M2) and 20 PRS (M3) to clinical baseline features (M1); evaluated under race-blind and race-aware specifications.

**Main Outcomes and Measures:** First inpatient ischemic stroke within 10 years; discrimination (area under the receiver operating characteristic curve [AUROC]) and calibration (observed-to-expected [O/E] ratio).

**Results:** In the AoU test partition (n = 6,998; 384 cases), M3 achieved AUROC 0.813 (95% CI, 0.788–0.837), outperforming the Revised Framingham Stroke Risk Profile (ΔAUROC 0.164) and Pooled Cohort Equations (ΔAUROC 0.181; both *P* < 0.001). Discrimination transferred to Bio*Me* (AUROC 0.745) but predictions were systematically high (aggregate O/E 0.12 vs. 1.00 in AoU), consistent with intercept-shift miscalibration; Bio*Me*-fit intercept recalibration restored calibration in African American and Hispanic but not European American participants. PRS contribution was significant only in the Bio*Me* Hispanic (ΔAUROC + 0.042; *P* = 0.003), with no significant within-stratum gain in five other cohort-by-race combinations. Adding self-reported race produced small gains when combined with PRS (Bio*Me* + 0.022, *P* = 0.034; AoU + 0.006, *P* = 0.052) but not when added without PRS.

**Conclusions and Relevance:** A machine-learning ensemble combining clinical, laboratory, and polygenic features outperformed traditional risk scores by 0.16–0.18 AUROC and retained discriminative validity in an ancestrally distinct external cohort but required site-specific recalibration of absolute risk. The marginal contribution of self-reported race overlapped with polygenic signal, supporting per-ancestry calibration over universal race-aware model deployment.

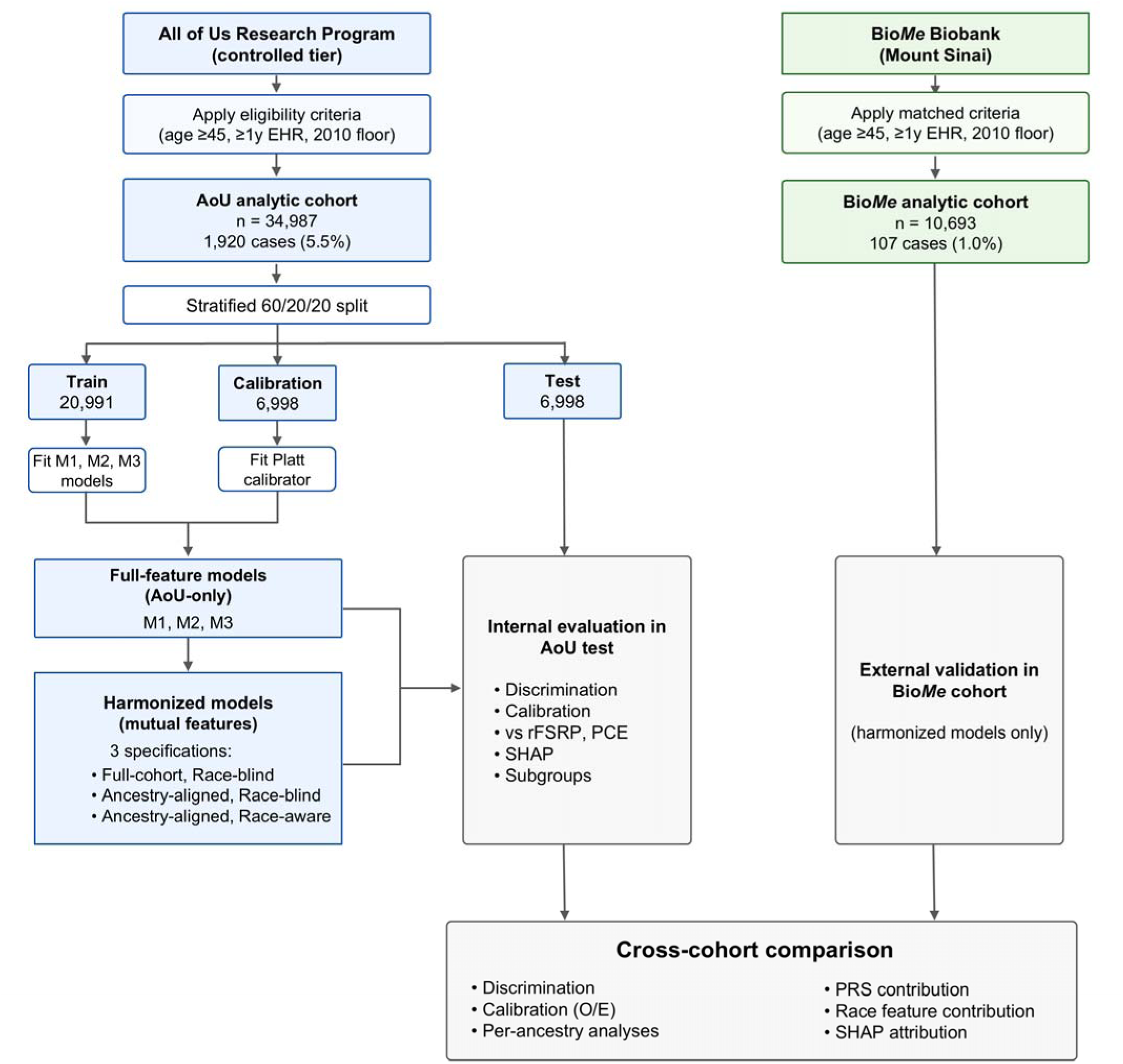

**Graphical Abstract:** Study design and analytic workflow. The All of Us (AoU) Research Program analytic cohort (n = 34,987; 1,920 incident ischemic strokes) was partitioned 60/20/20 into train, calibration, and held-out test sets. Two parallel model families were trained on the AoU train partition: full-feature models using all AoU features (evaluated only in AoU), and harmonized models using the intersection of AoU and Bio*Me* features (evaluated in both cohorts). Each family comprised three nested tiers (M1, clinical baseline; M2, M1 plus laboratory feature trajectories; M3, M2 plus 20 polygenic risk scores). Three specifications of the harmonized models addressed ancestry-related questions: full-cohort race-blind (primary cross-cohort analyses), ancestry-aligned race-blind, and ancestry-aligned race-aware. The Bio*Me* Biobank cohort at Mount Sinai (n = 10,693; 107 incident ischemic strokes) served as the external validation set under matched eligibility criteria and an identical inpatient ischemic stroke outcome definition. Internal evaluation in the AoU test partition included discrimination, calibration, comparison with the Revised Framingham Stroke Risk Profile (rFSRP) and Pooled Cohort Equations (PCE), Shapley additive explanations (SHAP) feature attribution, and subgroup analyses. Cross-cohort analyses evaluated discrimination, calibration (observed-to-expected [O/E] ratio), per-ancestry performance, domain-level SHAP attribution, and the marginal contributions of polygenic risk scores and self-reported race.

## Introduction

Ischemic stroke is a leading cause of death and acquired disability worldwide.^1^ Effective primary prevention requires accurate, equitable identification of individuals at elevated stroke risk so that preventive interventions, antihypertensive therapy, lipid-lowering treatment, antiplatelet therapy, and lifestyle modification, can be targeted to those most likely to benefit.^2^ Stroke incidence and outcomes vary substantially by race and ethnicity in the United States, with disproportionate burden among Black and Hispanic populations,^3,4^ making the equitable performance of risk-prediction tools across populations a clinical and methodological priority. Traditional risk scores such as the Framingham Stroke Risk Profile (rFSRP)^5^ and the Pooled Cohort Equations (PCE)^6^ have anchored clinical decision-making for two decades but were derived from cohorts that under-represent the United States diverse populations and exclude clinically meaningful electronic health record (EHR)-derived features such as laboratory feature trajectories and polygenic risk.

Machine-learning approaches offer an opportunity to leverage the rich clinical data, and a growing number of studies have applied them to stroke risk prediction.^7–9^ Yet important gaps remain before these models can be deployed equitably:^10^ many have been developed in single health systems with limited ancestral diversity, and few have been externally validated in cohorts with distinct ancestry compositions and clinical practice patterns. Beyond external validation, two questions are especially important for fair implementation: (1) whether self-reported race and ethnicity contributes meaningfully to model performance once other ancestry-correlated features are present, particularly in the context of recent efforts to remove race-based coefficients from clinical algorithms,^11,12^ and (2) whether polygenic risk scores (PRS), historically derived from primarily European-ancestry populations, provide consistent benefit across ancestries when added to EHR-based models.^13,14^

The All of Us (AoU) Research Program^15^ and the Bio*Me* Biobank^16^ together offer a deliberate design for addressing these questions: a national research cohort with ancestral diversity reflective of the contemporary U.S. population paired with an academic medical center biobank with distinct ancestry composition and clinical setting. Both cohorts include linked EHR and genetic data mapped to the Observational Medical Outcomes Partnership (OMOP) Common Data Model, enabling harmonized feature extraction and independent validation. Using AoU for model development and Bio*Me* for external validation, we developed a tiered machine-learning framework for 10-year incident ischemic stroke prediction. We assessed whether longitudinal laboratory features and PRS improved prediction beyond baseline clinical risk factors, and whether self-reported race contributed additional predictive information after accounting for ancestry-correlated clinical and genetic features. We compared performance with established traditional risk scores and evaluated discrimination, calibration, and clinical utility overall and across ancestry groups.

## Methods

We developed and externally validated machine-learning models for 10-year incident ischemic stroke prediction. Models were trained in AoU and externally validated in the Bio*Me* Biobank. Both cohorts contributed harmonized OMOP-structured EHR data. The study followed the Transparent Reporting of a multivariable prediction model for Individual Prognosis or Diagnosis + Artificial Intelligence (TRIPOD+AI) reporting guideline.

## Cohort construction and outcome

Cohort overviews are provided in Supplementary Methods 1. *AoU.* Participants were eligible if they were aged ≥45 years at baseline and had ≥1 year of EHR observation prior to baseline. The baseline date was the latest of (i) first EHR contact at age ≥45, (ii) first EHR contact plus 1 year, and (iii) January 1, 2010 (a calendar floor introduced to address severe sparsity and atypical control enrichment in pre-2010 records). The cohort was partitioned with a stratified 60/20/20 train/calibration/test split. Bio*Me.* External validation applied identical age, lookback, outcome, and calendar-floor criteria.

### Outcome

The outcome was incident inpatient ischemic stroke within 10 years of baseline. Cases were defined as the first occurrence of cerebral infarction (International Classification of Diseases [ICD], Ninth Revision codes 433.x1 or 434.x1; Tenth Revision codes I63.xx) during an inpatient encounter (Supplementary Table 1). Non-ischemic stroke phenotypes were excluded entirely; participants with only non-inpatient ischemic stroke records were removed. Controls required ≥10 years of follow-up. As a BioMe sensitivity analysis, we evaluated a relaxed outcome definition in which the first ischemic stroke ICD code during an inpatient or outpatient encounter was classified as an incident event; participants with only outpatient ischemic stroke records were retained as cases.

## Feature engineering

Feature engineering incorporated pre-baseline demographic, clinical (Supplementary Table 1), laboratory, medication, survey, anthropometric, and genetic data, with full details provided in Supplementary Methods 2. Twenty PRS from the Polygenic Score (PGS) Catalog^17^ (Supplementary Table 2) were computed from AoU whole-genome sequencing data using AoUPRS^18^ and from Bio*Me* genotyping array data imputed to TOPMed v2. Estimated glomerular filtration rate (eGFR) was derived using the Chronic Kidney Disease Epidemiology Collaboration (CKD-EPI) 2021 race-free equation. No post-baseline information entered any model. All features used in the models are listed in Supplementary Tables 3-4.

## Model architecture

Three nested model tiers reflect deployment contexts with increasing data demands: M1 (clinical baseline features), M2 (M1 + biomarker trajectories), and M3 (M2 + 20 PRS) (Supplementary Methods 2, Supplementary Table 4). All tiers used extreme gradient boosting (XGBoost) decision trees,^19^ which route missing values to the loss-minimizing branch at each split without imputation. Hyperparameters were tuned with Optuna^20^ across 100 trials of 5-fold stratified cross-validation on the train partition, optimizing average precision (Supplementary Table 5). Models were refit on the full train partition with selected hyperparameters and frozen. Platt scaling^21^ was fit once on the AoU calibration partition and applied unchanged to both the AoU test partition and the Bio*Me* validation cohort. As a post-hoc sensitivity analysis of the cross-cohort calibration drift, an intercept-only logistic recalibration was additionally fit on a stratified 50% partition of the Bio*Me* cohort and evaluated in the held-out 50%. The probability threshold achieving 90% specificity was derived independently within each cohort.

## Analytic framework

Two parallel model families were trained. The *full-feature* models used the full AoU feature set with self-reported race indicators. Because several features (e.g., AoU survey instruments) have no Bio*Me* equivalent, these models could be evaluated only in AoU; they supported the internal validation analyses, comparison with the rFSRP and PCE within eligible subsets (eligibility criteria detailed in Supplementary Methods 3), Shapley additive explanations (SHAP) feature attribution^22,23^, and subgroup performance by sex, age, and prevalent diabetes.

The *harmonized* models used the intersection of AoU and Bio*Me* feature sets, so that a single AoU-trained model could be evaluated in both the AoU held-out test partition and the Bio*Me* validation cohort without imputation. Three specifications of the harmonized models were trained: (1) *full cohort, race-blind* — entire AoU cohort with race excluded, providing the primary cross-cohort and ancestry-stratified analyses; (2) *ancestry-aligned, race-blind* — restricted to participants self-identifying as European American, African American, or Hispanic (the three groups with adequate counts in both cohorts), race excluded; and (3) *ancestry-aligned, race-aware* — the same restricted cohort with race indicators added. The (3) vs (2) contrast isolates the marginal contribution of self-reported race (see Supplementary Methods 2).

## Statistical analysis

Discrimination was assessed using the area under the receiver operating characteristic curve (AUROC) and the area under the precision-recall curve (AUPRC); calibration using the observed-to-expected (O/E) event ratio, intercept, and slope; and threshold-dependent metrics were reported at the per-cohort 90%-specificity threshold (full metric definitions in Supplementary Methods 4). Confidence intervals were 2,000-bootstrap percentile intervals. Pairwise AUROC comparisons across nested tiers used the DeLong test^24^ on rows aligned by participant identifier; pairwise AUPRC comparisons used a paired bootstrap with two-sided p-values. Per-ancestry analyses (AUROC, calibration, paired M2 vs M3 ΔAUROC) were performed within strata of the race-blind full-cohort model on the held-out test/validation set, without ancestry-specific retraining. Treatment-stratified O/E ratios were computed within Bio*Me* by ancestry, with treatment defined as any-ever baseline exposure to statins, angiotensin-converting enzyme inhibitors (ACE inhibitors), angiotensin II receptor blockers (ARBs), antiplatelets, warfarin, factor Xa inhibitors, or direct thrombin inhibitors. Domain-level SHAP attribution was computed in both the AoU test partition and Bio*Me* validation cohort for the harmonized full-cohort race-blind M3 model, with mean absolute SHAP values aggregated within nine clinical feature domains and expressed as a percentage of the per-cohort total. Analyses used Python 3.10 with XGBoost 3.0.0, scikit-learn 1.6.0, statsmodels 0.14.4, and Optuna 4.5.0.

## Results

### Cohort characteristics

The AoU development cohort comprised 34,987 participants with 1,920 (5.5%) incident ischemic strokes over 10 years; the held-out test partition included 6,998 participants and 384 cases. The Bio*Me* external validation cohort comprised 10,693 participants with 107 (1.0%) incident strokes (Table 1). Two patterns distinguished the cohorts: Bio*Me* contained higher proportions of Hispanic (35% vs 12%) and Black or African American (22% vs 19%) participants than AoU, and Bio*Me* controls had 5- to 20-fold higher statin, antiplatelet, and ACE inhibitor exposure than AoU controls (Table 1).

**Table 1.** Baseline Characteristics of the Ischemic Stroke Risk Prediction Cohorts.

| Characteristic | AoU Controls<br>N=33,067 | AoU Cases<br>N=1,920 | BioMe Controls<br>N=10,586 | BioMe Cases<br>N=107 |
| --- | --- | --- | --- | --- |
| **Demographics** |  |  |  |  |
| Age at baseline, mean (SD), y | 54.37 (8.84) | 58.23 (10.86) | 58.54 (9.12) | 63.42 (9.59) |
| Male sex, No. (%) | 12,962 (39.2) | 945 (49.2) | 3,808 (37.2) | 38 (36.2) |
| **Race and ethnicity, No. (%)** |  |  |  |  |
| White | 21,295 (64.4) | 918 (47.8) | 2,964 (29.0) | 13 (12.4) |
| Black / African American | 6,448 (19.5) | 611 (31.8) | 2,246 (22.0) | 28 (26.7) |
| Hispanic | 3,968 (12.0) | 309 (16.1) | 3,578 (35.0) | 47 (44.8) |
| Asian | 893 (2.7) | 35 (1.8) | NA | NA |
| East Asian | NA | NA | 243 (2.4) | 0 (0.0) |
| South Asian | NA | NA | 150 (1.5) | 3 (2.9) |
| Other, multiple, or unknown | 2,745 (8.3) | 200 (10.4) | 1,051 (10.3) | 14 (13.3) |
| **Cardiovascular and cerebrovascular history, No. (%)** |  |  |  |  |
| Hypertension | 7,209 (21.8) | 979 (51.0) | 3,899 (36.8) | 56 (52.3) |
| Atrial fibrillation | 364 (1.1) | 109 (5.7) | 207 (2.0) | 9 (8.4) |
| Coronary artery disease | 860 (2.6) | 255 (13.3) | 680 (6.4) | 18 (16.8) |
| Heart failure | 430 (1.3) | 182 (9.5) | 220 (2.1) | 13 (12.1) |
| Peripheral arterial disease | 66 (0.2) | 23 (1.2) | 192 (1.8) | 7 (6.5) |
| Carotid artery disease | 66 (0.2) | 44 (2.3) | 49 (0.5) | 1 (0.9) |
| Transient ischemic attack | 66 (0.2) | 42 (2.2) | 85 (0.8) | 2 (1.9) |
| **Metabolic, kidney, and respiratory history, No. (%)** |  |  |  |  |
| Type 2 diabetes | 2,645 (8.0) | 449 (23.4) | 1,762 (16.6) | 35 (32.7) |
| Chronic kidney disease | 463 (1.4) | 167 (8.7) | 473 (4.5) | 13 (12.1) |
| Dyslipidemia | 6,514 (19.7) | 616 (32.1) | 3,416 (32.3) | 51 (47.7) |
| Obesity | 2,877 (8.7) | 349 (18.2) | 1,150 (10.9) | 10 (9.3) |
| Sleep apnea | 1,455 (4.4) | 173 (9.0) | 424 (4.0) | 7 (6.5) |
| Chronic obstructive pulmonary disease | 562 (1.7) | 113 (5.9) | 213 (2.0) | 8 (7.5) |
| **Clinical measurements, mean (SD)** |  |  |  |  |
| Systolic blood pressure, mm Hg | 125.28 (16.72) | 134.61 (21.49) | 127.26 (17.87) | 132.40 (19.19) |
| Diastolic blood pressure, mm Hg | 76.27 (10.43) | 79.12 (12.86) | 75.45 (10.47) | 75.66 (12.40) |
| HbA1c, % | 6.27 (1.49) | 7.00 (2.04) | 6.38 (1.41) | 6.90 (2.04) |

| Characteristic | AoU Controls<br>N=33,067 | AoU Cases<br>N=1,920 | BioMe Controls<br>N=10,586 | BioMe Cases<br>N=107 |
| --- | --- | --- | --- | --- |
| eGFR, mL/min/1.73 m<sup>2</sup> | 89.10 (18.70) | 81.69 (24.13) | 79.01 (19.59) | 70.88 (27.52) |
| LDL cholesterol, mg/dL | 109.69 (32.75) | 102.29 (39.79) | 102.56 (33.55) | 95.79 (35.83) |
| HDL cholesterol, mg/dL | 52.57 (17.05) | 48.36 (17.60) | 56.34 (17.98) | 50.88 (17.16) |
| **Medication exposure, No. (%)** |  |  |  |  |
| Antiplatelet exposure | 860 (2.6) | 33 (1.7) | 1,506 (14.2) | 38 (35.5) |
| Factor Xa inhibitor exposure | 0 (0.0) | 10 (0.5) | 8 (0.1) | 1 (0.9) |
| Direct thrombin inhibitor exposure | 0 (0.0) | 2 (0.1) | 19 (0.2) | 1 (0.9) |
| Heparin exposure | 496 (1.5) | 21 (1.1) | 140 (1.3) | 5 (4.7) |
| Warfarin exposure | 165 (0.5) | 25 (1.3) | 212 (2.0) | 5 (4.7) |
| Statin exposure | 298 (0.9) | 21 (1.1) | 2,155 (20.4) | 41 (38.3) |
| ACE inhibitor exposure | 463 (1.4) | 83 (4.3) | 1,696 (16.0) | 28 (26.2) |
| ARB exposure | 132 (0.4) | 27 (1.4) | 797 (7.5) | 10 (9.3) |

Values are presented as mean (SD) or No. (%). Race and ethnicity categories may overlap; percentages do not sum to 100. Abbreviations: ACE, angiotensin-converting enzyme; ARB, angiotensin receptor blocker; eGFR, estimated glomerular filtration rate; HbA1c, hemoglobin A1c; HDL, high-density lipoprotein; LDL, low-density lipoprotein; NA, not available or not applicable.

### AoU internal validation

In the AoU held-out test partition (n = 6,998; 384 cases), the full-feature M3 model achieved an AUROC of 0.813 (95% CI, 0.788–0.837) and an AUPRC of 0.380 (0.330–0.429) (Figures 1a-1b). Adding laboratory trajectory features (M2 vs M1) significantly improved discrimination (ΔAUROC 0.020, DeLong *P* = 0.016; ΔAUPRC 0.068, paired bootstrap *P* < 0.001), while the further increment from polygenic risk scores (M3 vs M2) was not statistically significant (ΔAUROC 0.004, *P* = 0.253; ΔAUPRC 0.009, *P* = 0.090) (Supplementary Table 6).

**Figure 1.**
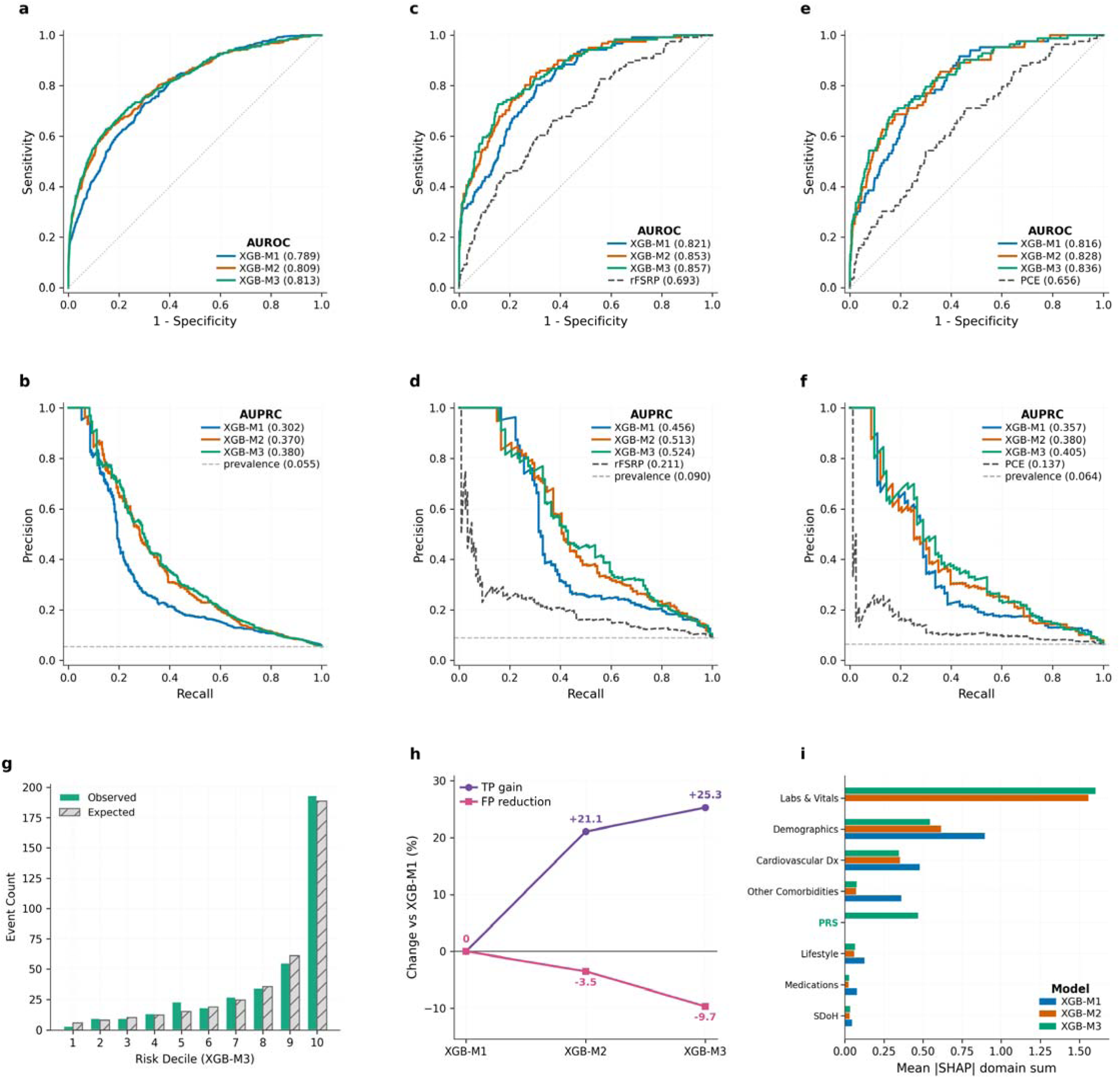
Internal validation of stroke risk models in the All of Us (AoU) test partition. **a–b**, Receiver operating characteristic **(a)** and precision-recall **(b)** curves for the three nested model tiers (M1, M2, M3) in the held-out AoU test set (n = 6,998; 384 incident stroke cases over 10 years; prevalence 5.49%). The dashed line in b indicates the case prevalence (precision-recall baseline). **c–d**, Same curves restricted to the rFSRP-eligible subset (n = 1,338; 121 cases; prevalence 9.04%), with rFSRP shown for direct comparison. **e–f**, Same curves restricted to the PCE-eligible subset (n = 1,287; 83 cases; prevalence 6.45%), with PCE shown for direct comparison. **g**, Calibration of M3 in the AoU test set: observed (solid) versus expected (hatched) event counts within each decile of M3-predicted risk. **h**, Change in true-positive captures (TP gain) and false-positive volume (FP reduction) for M2 and M3 relative to M1 at a fixed 90%-specificity operating policy applied to the AoU test set. **i**, Mean absolute SHAP value summed within each feature domain, plotted for all three model tiers and ranked top-to-bottom by M3 contribution. The Family History domain (zero contribution across all tiers) is omitted from display.

Within the subsets eligible for traditional risk scores, M3 substantially outperformed the rFSRP (rFSRP-eligible subset: n = 1,338; 121 cases; M3 AUROC 0.857 vs. rFSRP 0.693; ΔAUROC 0.164, *P* < 0.001; Figures 1c - 1d) and the PCE (PCE-eligible subset: n = 1,287; 83 cases; M3 AUROC 0.836 vs. PCE 0.656; ΔAUROC 0.181, *P* < 0.001; Figure 1e - 1f).

### AoU calibration, operating point, and feature attribution

Calibrated M3 probabilities aligned closely with observed event counts in the AoU test set (Figure 1g), with the highest-risk decile capturing approximately half of all incident strokes. At the 90%-specificity threshold, M3 captured 54% of incident strokes, compared with 43% for M1 (positive predictive value [PPV], 25% vs. 20%; Figure 1h; Supplementary Table 6), corresponding to 25% more true positives and 10% fewer false positives under the same operating policy. Domain-level SHAP attribution shifted substantially across tiers (Figure 1i): M1 was dominated by demographics and cardiovascular history; M2 and M3 by laboratory and vital-sign features; and, in M3, PRS emerged as the third-largest domain after laboratory/vital-sign features and demographics.

AUC values are shown in panel legends. AUPRC = area under the precision-recall curve; AUROC = area under the receiver operating characteristic curve; FP = false positive; M1 = clinical baseline model (demographics, surveys, comorbidities, vitals, anthropometrics, medications); M2 = M1 plus laboratory trajectory features; M3 = M2 plus polygenic risk scores; PCE = Pooled Cohort Equations; PRS = polygenic risk score; rFSRP = Revised Framingham Stroke Risk Profile; SHAP = SHapley Additive exPlanations; TP = true positive.

### Subgroup performance in AoU

M3 performance within sex, age, and prevalent-diabetes strata (Figure 2) showed preserved discrimination across all strata (AUROC 0.769–0.828), with essentially identical discrimination between younger (45–64) and older (≥65) participants despite a more than 2-fold difference in event rate. Calibration was well-aligned in five of six strata (O/E 0.97–1.05); the exception was prevalent diabetes (n = 594; 96 cases; event rate 16.2%), in which the model modestly underpredicted risk (O/E 1.20 [1.01–1.40]; Figure 2k). Decision curve analysis showed positive net benefit of M3 over both treat-all and treat-none strategies across the screening-relevant threshold range (1–30%) in all subgroups (Figure 2d, 2h, 2l).

**Figure 2.**
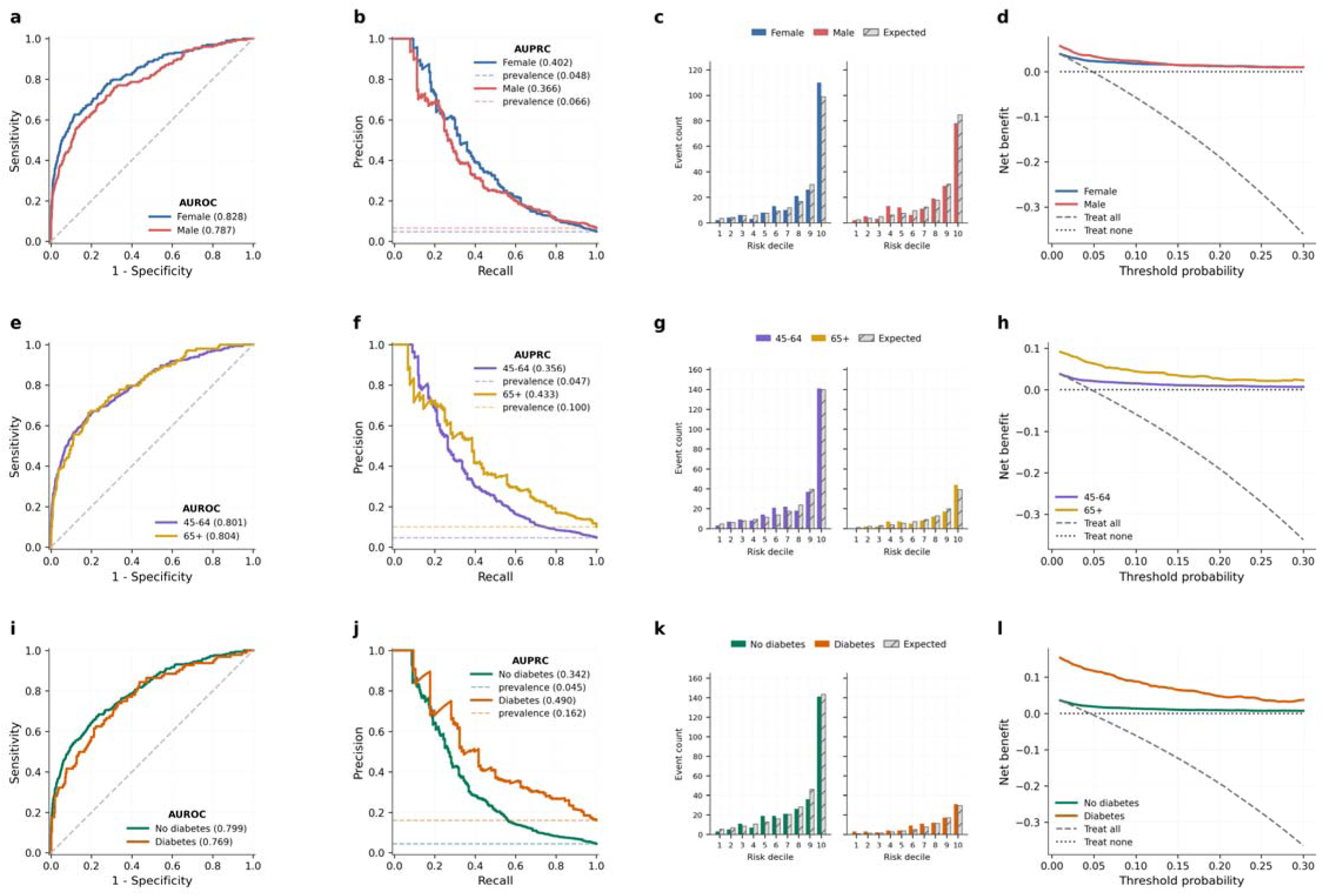
Subgroup performance of the M3 model in the All of Us (AoU) test partition. Performance of the full-feature M3 model (clinical features + laboratory trajectories + polygenic risk scores, full AoU feature set with race indicators) evaluated within strata of sex (a–d), age group (e–h), and prevalent diabetes (i–l) in the held-out AoU test set (n = 6,998; 384 incident stroke cases). Each row presents four panels for one subgroup variable: receiver operating characteristic curves with AUROC values in the legend **(a, e, i)**; precision-recall curves with AUPRC values in the legend, with dashed lines indicating subgroup-specific case prevalence **(b, f, j)**; calibration plots showing observed (solid colored bars) versus expected (gray hatched bars) event counts within deciles of M3-predicted risk, with side-by-side sub-panels for the two strata of each subgroup variable **(c, g, k)**; and decision curve analysis comparing M3 net benefit (solid colored lines) with treat-all (dashed gray) and treat-none (dotted, at zero) strategies across the threshold-probability range relevant to stroke screening (1–30%) **(d, h, l)**. AUPRC = area under the precision-recall curve; AUROC = area under the receiver operating characteristic curve; M3 = clinical baseline plus laboratory trajectories plus polygenic risk scores; O/E = observed-to-expected event ratio.

### External validation in Bio*Me*

We next applied the harmonized models to the Bio*Me* cohort (n = 10,693; 107 incident ischemic strokes). The full-cohort race-blind M3 model achieved an AUROC of 0.745 (95% CI, 0.699–0.787), an attenuation of approximately 0.06 AUROC from the same harmonized model applied to the AoU test partition (M3 AUROC 0.805; Figure 3e, f; Supplementary Table 7). The M2 vs M1 increment was substantial (Bio*Me* ΔAUROC 0.094), while the M3 vs M2 increment was small and not statistically significant (Bio*Me* ΔAUROC 0.014, DeLong *P* = 0.098; Supplementary Table 8). A sensitivity analysis using the relaxed outcome definition captured 849 incident events in Bio*Me* (vs. 107 under the strict definition); M3 AUROC was lower across all three specifications (0.646–0.674 vs 0.739–0.761; Supplementary Table 9).

**Figure 3.**
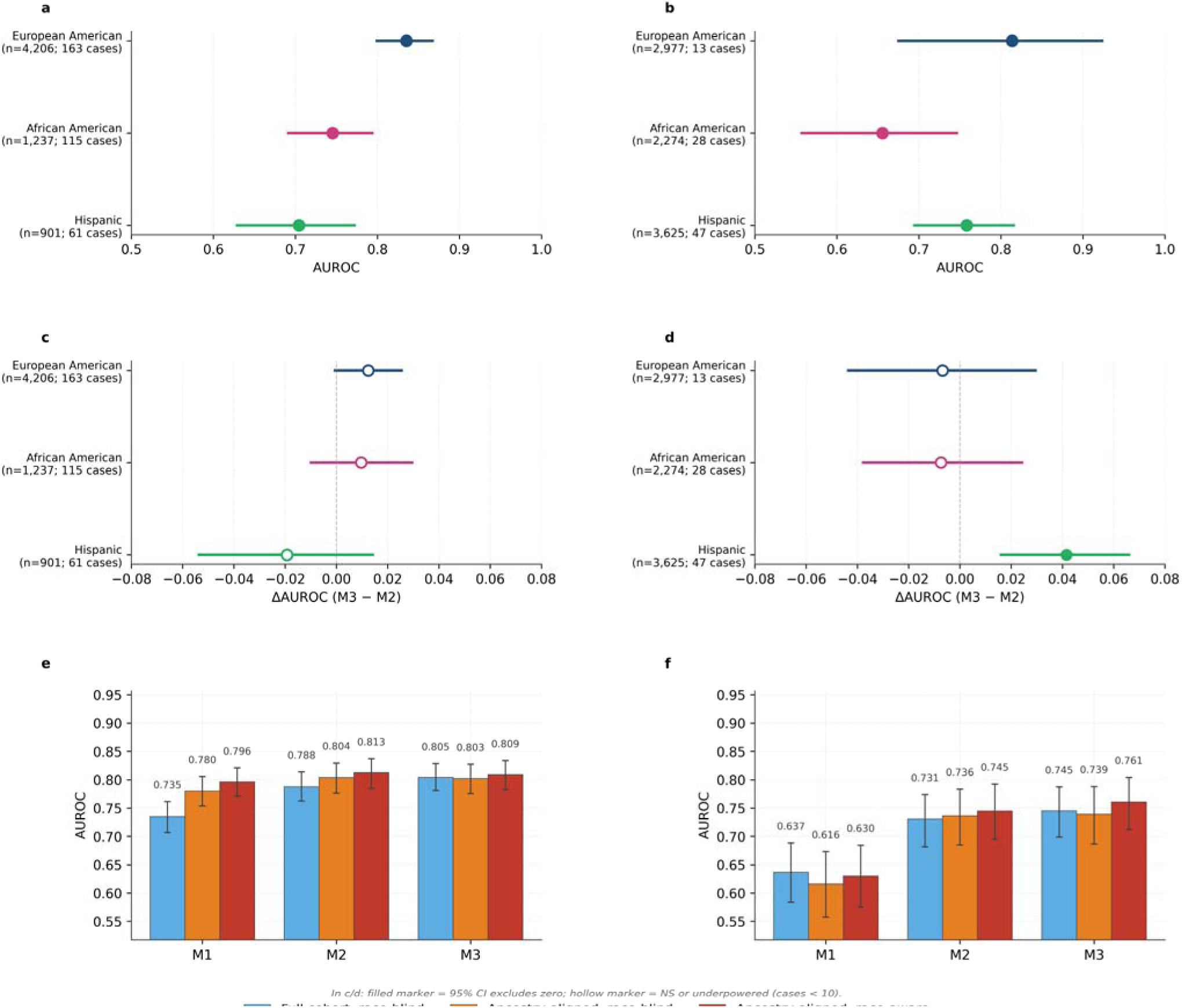
Ancestry-stratified discrimination and polygenic risk score contribution of the M3 stroke prediction model in the AoU test set and the BioMe external validation cohort. **a–b**, M3 AUROC by self-reported ancestry under the full-cohort race-blind specification in the AoU held-out test set (a; n = 6,998; 384 incident ischemic strokes) and the BioMe validation cohort (b; n = 10,693; 107 incident ischemic strokes). Markers indicate point estimates; horizontal bars are 95% percentile-bootstrap CIs (2,000 resamples). The vertical dashed line marks AUROC = 0.5. **c–d**, Paired within-ancestry contribution of polygenic risk scores, computed as ΔAUROC for M3 versus M2 in the same race-blind full-cohort specification, in AoU (c) and BioMe (d). Predictions for M2 and M3 are aligned at the participant level so that the same individuals contribute to both estimates. Filled markers indicate that the 95% CI excludes zero; hollow markers indicate non-significant comparisons. The vertical dashed line marks ΔAUROC = 0. **e–f**, M3 discrimination AUROC by tier (M1, M2, M3) and model specification (full cohort, race-blind; ancestry-aligned cohort, race-blind; ancestry-aligned cohort, race-aware) in AoU (e) and BioMe (f). Error bars are 95% percentile-bootstrap CIs. Y-axes are harmonized across cohorts to enable direct cross-site comparison. The BioMe outcome is defined as a first inpatient ischemic stroke within 10 years of the landmark date, matching the AoU phenotype; per-stratum numerical results including these strata are reported in Supplementary Table 10. AUROC = area under the receiver operating characteristic curve; M1 = baseline clinical features; M2 = M1 + laboratory feature trajectories; M3 = M2 + polygenic risk scores.

Discrimination transferred, but Bio*Me* risk predictions were systematically high (aggregate O/E ratio 0.12 in Bio*Me* vs 1.00 in AoU; Supplementary Figure 1 and Supplementary Table 7). Calibration slopes remained near 1 across Bio*Me* ancestry strata, consistent with intercept-shift miscalibration. Intercept-only recalibration shifted O/E toward unity in African American (1.05) and Hispanic (0.86) participants, with residual under-correction in European American participants (0.43; Supplementary Figure 2). Despite the marked cohort exposure gap (Table 1), treated and untreated Bio*Me* participants exhibited similar O/E values within each ancestry stratum (Supplementary Figure 2), indicating that the calibration miss was not primarily explained by baseline cardiovascular medication exposure.

### Ancestry-stratified performance and contribution of polygenic risk scores

Per-ancestry analyses were restricted to European American, African American, and Hispanic participants (≥10 cases per cohort). M3 discrimination varied by self-reported ancestry in both cohorts (Figure 3a, b). In AoU, AUROC was highest in European American participants (0.835 [0.798–0.869]), lower in African American (0.745 [0.690–0.795]) and Hispanic participants (0.704 [0.627–0.774]). In Bio*Me*, AUROC was preserved in European American (0.814 [0.673–0.925]) and Hispanic participants (0.758 [0.693–0.817]) but lower in African American participants (0.655 [0.556–0.748]; Supplementary Table 10). The within-stratum contribution of PRS (paired ΔAUROC for M3 vs M2 within each ancestry; Figure 3c,d) was small and not statistically significant in five of the six combinations of cohort and ancestry (point estimates −0.019 to +0.012), despite a small but significant pooled M3 vs M2 increment in the AoU harmonized model (ΔAUROC 0.017, *P* = 0.004; Supplementary Table 8), consistent with limited within-stratum power. The exception was the Bio*Me* Hispanic stratum, in which PRS produced a significant discrimination gain (ΔAUROC +0.042 [+0.016, +0.067]; paired bootstrap *P* = 0.003; Supplementary Table 11). Consistent with this finding, domain-level SHAP attribution showed higher PRS contribution in Bio*Me* than AoU across all ancestry strata, with the largest cross-cohort difference observed in Hispanic participants (19.8% vs 14.8% of total mean |SHAP|; Supplementary Figure 3). Adding self-reported race as a model feature produced small AUROC gains in both cohorts: ΔAUROC 0.006 (95% CI, -0.001 to 0.013; *P* = 0.053) in AoU and 0.022 (0.001 to 0.044; *P* = 0.035) in Bio*Me* (Figure 3e, f; Supplementary Table 12). The corresponding race-aware comparison in the M2 specification (without PRS) was not statistically significant in either cohort (AoU ΔAUROC 0.008, *P* = 0.099; Bio*Me* ΔAUROC 0.009, *P* = 0.404; Supplementary Table 12).

## Discussion

In two ancestrally diverse cohorts, a tiered XGBoost model for 10-year ischemic stroke risk substantially outperformed traditional risk scores in AoU and retained discrimination upon external validation in the Bio*Me* cohort. Laboratory feature trajectories — summarizing means, variability, slopes, and recency of repeated biomarker measurements — provided the dominant incremental contribution to discrimination; PRS produced small and ancestry-heterogeneous additional gains. Self-reported race contributed a small but statistically detectable discrimination improvement in the more ancestrally diverse Bio*Me* cohort, with a borderline equivalent in AoU; this gain was modest in magnitude and appeared to reflect ancestry-related signal partially overlapping with that carried by PRS.

Prior machine-learning approaches to primary-prevention ischemic stroke risk prediction have generally reported AUROCs of approximately 0.70 to 0.80,^25–28^ comparable to or modestly below the 0.81 reported here. These studies have primarily used point-in-time clinical features rather than repeated-measures laboratory trajectories, have rarely integrated polygenic information^29^, and have not consistently been externally validated across ancestrally distinct cohorts; few have reported calibration alongside discrimination,^25^ which limits assessment of cross-cohort transferability. Prior work has separately addressed repeated-measures biomarkers for stroke prediction,^30^ longitudinal EHR features for CVD prediction,^31^ and their integration with genetic data at single sites,^32^ but their combination for ischemic stroke with external validation across ancestrally distinct cohorts has not been reported. Studies reporting higher AUROCs (≥0.85) in stroke prediction have typically included prior stroke history as a feature or used short prediction windows (e.g., 1 year), reflecting recurrent or near-term events rather than 10-year incident first-stroke risk.^33^. A 2010 baseline floor and 10-year follow-up requirement reduced bias from sparse historical documentation and unequal outcome ascertainment.

Discrimination transferred to Bio*Me*, but absolute risk estimates were systematically too high, a pattern commonly observed when prediction models are externally validated in populations differing from the development cohort.^34,35^ Calibration slopes remained near 1, consistent with intercept-shift miscalibration. Intercept-only recalibration restored calibration in African American and Hispanic participants but left residual under-correction in European American participants, suggesting per-ancestry recalibration may be more appropriate than a single global correction. Treatment-stratified analysis showed similar O/E values in treated and untreated Bio*Me* participants within each ancestry, indicating the cohort medication-exposure gap does not explain the calibration miss.

In the AoU test set PRS was the third most important feature in the full model (M3) ahead of lifestyle, medication and SDOH features, however in BioMe PRS contribution was statistically significant only in Hispanic participants (ΔAUROC +0.042). The BioMe analysis could have been limited by smaller sample sizes in the individual race stratum, but these results are not inconsistent with the cardiometabolic literature.^36^ Other studies in CVDs have shown PRS to contribute meaningfully to the prediction of events using machine learning model approaches in the presence of traditional risk factors but the degree of improvement is variable with several studies reporting modest gains.. Self-reported race added marginal improvement only when combined with PRS, consistent with race and polygenic features carrying overlapping ancestry-correlated signal.

The rFSRP AUROC of 0.693 here is consistent with external validations reporting C-statistics of 0.71–0.74.^37,38^ PCE’s lower discrimination (0.656) likely reflects its derivation for the atherosclerotic cardiovascular disease (ASCVD) composite outcome^6^ rather than for stroke alone, consistent with attenuated PCE performance in modern diverse cohorts.^39^ Prior validations of both scores have either imputed missing inputs^38^ or excluded predictors where EHR missingness was not at random;^40^ XGBoost’s native missing-value handling avoids both issues.

Some limitations merit consideration. First, both cohorts are derived from EHR data, so feature ascertainment and outcome capture are subject to documentation patterns that may differ across health systems. Differential cross-system EHR linkage between the multi-site AoU cohort and the single-system Bio*Me* cohort likely contributes to the lower Bio*Me* event rate. A sensitivity analysis using a relaxed outpatient-inclusive outcome captured eight-fold more events but with consistently lower discrimination (Supplementary Table 9), consistent with the lower PPV of outpatient ICD codes for incident stroke;^41–43^ we retained the strict definition as primary. Second, PRS were derived using PGS Catalog scores predominantly developed in European-ancestry populations; we did not evaluate ancestry-specific PRS construction. Third, although Bio*Me*’s diversity exceeds that of many single-site validation cohorts, broader validation of the model and recalibration approach across additional health systems and geographies is needed.

A nested machine-learning ensemble combining clinical, laboratory-trajectory, and polygenic features outperformed traditional risk scores and retained discriminative validity in an external academic medical center cohort, with positive net benefit across the screening-relevant threshold range. The cross-cohort calibration drift in Bio*Me* indicates that deployment of EHR-trained risk models should include local calibration assessment and, when needed, intercept-level recalibration. The small marginal contributions of self-reported race observed here support pursuing equitable performance through ancestry-stratified calibration assessment rather than universal race-aware model deployment.

## Data Availability

The data that supports the findings of this study are available from the All of Us Research Program and the Mount Sinai BioMe Biobank, but restrictions apply to the availability of these data, which were used under data access agreements for the current study and so are not publicly available. All of Us data are available to authorized researchers through the All of Us Researcher Workbench (https://www.researchallofus.org) with permission of the All of Us Research Program. BioMe data is available through approved institutional processes with permission of the Mount Sinai BioMe Biobank.

## Declarations

### Ethics approval and consent to participate

This study used de-identified data from the All of Us Research Program and the Mount Sinai Bio*Me* Biobank. The All of Us Research Program operates under a centralized Institutional Review Board that follows the regulations and guidance of the Office for Human Research Protections (OHRP), and all participants provided informed consent for their data to be used in approved research at the time of enrollment. Access to All of Us data was conducted through an All of Us-participating institution under the program’s established data access framework. The Bio*Me* Biobank operates under the Mount Sinai Institutional Review Board, and all participants provided written informed consent at enrollment. Leveraging these datasets for the present study was approved by the University of Alabama at Birmingham Institutional Review Board (IRB-300006721). The study was conducted in accordance with the principles of the Declaration of Helsinki.

### Consent for publication

Not applicable.

### Availability of data and materials

The data that supports the findings of this study are available from the All of Us Research Program and the Mount Sinai Bio*Me* Biobank, but restrictions apply to the availability of these data, which were used under data access agreements for the current study and so are not publicly available. All of Us data are available to authorized researchers through the All of Us Researcher Workbench (https://www.researchallofus.org) with permission of the All of Us Research Program. Bio*Me* data is available through approved institutional processes with permission of the Mount Sinai Bio*Me* Biobank.

## Author contributions

A.K. conceived the study, designed the analytical framework, performed the analyses, and drafted the manuscript. Z.W. conducted the Bio*Me* data curation and external validation analyses, contributed to methodological development, and reviewed and revised the manuscript. V.S. contributed to methodological discussions and data curation strategy. H.K.T. contributed to methodological guidance, interpretation of results, and critical review and revision of the manuscript. R.L. contributed to the design and oversight of the Bio*Me* component and to manuscript review and revision. N.L. contributed to study oversight, interpretation of findings, and manuscript review and revision. M.R.I. supervised the study, contributed to study design, methodological guidance, and interpretation of findings, and critically revised the manuscript. All authors reviewed and approved the final manuscript.

## Acknowledgments

We gratefully acknowledge All of Us participants for their contributions, without whom this research would not have been possible. We also thank the National Institutes of Health’s All of Us Research Program for making available the participant data examined in this study.

The Mount Sinai Bio*Me* Biobank is supported by The Andrea and Charles Bronfman Philanthropies and by Federal funds from the NIH (U01HG00638001; U01HG007417; X01HL134588). We thank all participants and all our recruiters who have assisted and continue to assist in data collection and management. Furthermore, analyses were in part supported through the computational and data resources and staff expertise provided by Scientific Computing and Data at the Icahn School of Medicine at Mount Sinai and by the Clinical and Translational Science Awards (CTSA) grant UL1TR004419 from the National Center for Advancing Translational Sciences. Research reported in this publication was also supported by the Office of Research Infrastructure of the National Institutes of Health under award number S10OD026880 and S10OD030463. The content is solely the responsibility of the authors and does not necessarily represent the official views of the National Institutes of Health.

## Supplementary information

Supplementary information is available for this paper.

