## Supplementary Method, Supplementary Figure for "Development and External Validation of a Machine Learning Model for 10-Year Ischemic Stroke Risk Prediction in Diverse Populations"

**Supplementary Methods 1. AoU and Bio*Me* cohort descriptions.**

*All of Us Research Program (AoU).* AoU is a national longitudinal research program initiated by the National Institutes of Health to enroll and follow a diverse research cohort of at least one million U.S. participants.¹⁴ AoU enrolls broadly across U.S. health-provider organizations, with deliberate over-recruitment of populations historically underrepresented in biomedical research — including racial and ethnic minorities, individuals from rural and lower-income settings, and individuals with disabilities. Participants contribute electronic health record (EHR) data linked across all participating care sites, survey responses, physical measurements, biospecimens, and (for a subset of participants) short-read whole-genome sequencing data. The present analysis used the AoU Curated Data Repository (CDR) v8 controlled-tier release, accessed through the AoU Researcher Workbench.

Bio*Me* *Biobank*. The Bio*Me* Biobank is an ongoing, prospective, hospital- and outpatient-based population research program operated by The Charles Bronfman Institute for Personalized Medicine at Mount Sinai. Bio*Me* has enrolled over 60,000 participants since September 2007. This EHR-linked biorepository enrolls participants non-selectively from the Mount Sinai Health System, which serves a diverse group of communities across the greater New York City area. At enrollment, participants consent to link their DNA and plasma samples to their EHR; this is further complemented by a questionnaire on demographic and lifestyle factors. Participants were aged between 18 and 89+ years old at recruitment, and European American, African American, and Hispanic ancestries are well represented.¹⁵ Genotyping was performed on Illumina Global Screening Array (GSA) and Global Diversity Array (GDA) platforms; variants were imputed to the TOPMed reference panel, version 2, using the TOPMed Imputation Server.

**Supplementary Methods 2. Feature engineering.**

*Pre-baseline windowing.* For every participant, only data recorded prior to the baseline date were used in feature construction; measurements on or after the baseline date were strictly excluded to prevent information leakage. The same windowing rule was applied to laboratory values, vital signs, comorbidity codes, medication exposures, and survey responses.

*EHR-derived comorbidity features.* Pre-baseline comorbidity features were derived from electronic health record diagnoses using curated Systematized Nomenclature of Medicine — Clinical Terms (SNOMED CT) ancestor concepts. Each comorbidity was defined by a clinically relevant SNOMED CT ancestor concept, and participants were assigned a binary indicator if any descendant diagnosis code mapped to that ancestor concept before baseline. This approach enabled standardized capture of disease histories across related diagnostic codes. The full list of comorbidity definitions and corresponding SNOMED CT ancestor concepts is provided in Supplementary Table 1.

*Longitudinal trajectory features.* Trajectory features summarized longitudinal laboratory values and vital signs to capture both biological evolution and patterns of clinical monitoring over time. For each laboratory test and vital sign, all measurements recorded prior to the baseline date were included.

For each measurement series, trajectory features captured complementary aspects of clinical course: summary measures of central tendency and variability (mean, standard deviation, coefficient of variation), directional trend (slope), and recency of measurement. Slopes were estimated at the individual level using ordinary least squares regression of laboratory values on time (years since first measurement), yielding units per year. Slopes were computed only when at least three measurements were available; otherwise, slope features were set to missing. Recency features were defined as the elapsed time between the most recent measurement and the baseline date, capturing the intensity and continuity of longitudinal monitoring within each clinical domain.

Clinically interpretable abnormality features were derived for selected laboratory tests, including indicators of whether values crossed clinically relevant thresholds and the proportion of measurements that were abnormal over time. The ever-abnormal flag distinguished between three states: 1 (any abnormal value observed), 0 (tested but never abnormal), and missing (never tested), thereby separating clinical absence of measurement from documented normal results.

Together, these trajectory-based features encode both biological instability and clinical observability, distinguishing individuals with sparse or infrequent data from those with actively monitored and evolving disease patterns.

*Cardiovascular and metabolic medication classes.* Pre-baseline medication exposure was identified through drug-name string matching against the Observational Medical Outcomes Partnership (OMOP) Common Data Model drug exposure table within each cohort, using a curated list of 29 cardiovascular and metabolic medication classes. Classes evaluated covered five therapeutic categories: (i) anticoagulants — warfarin, direct thrombin inhibitors, factor Xa inhibitors, and heparins; (ii) antiplatelets; (iii) lipid-lowering agents — statins, fibrates, ezetimibe, and PCSK9 inhibitors; (iv) antihypertensives — angiotensin-converting enzyme (ACE) inhibitors, angiotensin II receptor blockers (ARBs), beta blockers, calcium channel blockers, alpha blockers, thiazide diuretics, loop diuretics, potassium-sparing diuretics, mineralocorticoid receptor antagonists, central-acting agents, and direct vasodilators; and (v) antidiabetic agents — metformin, sulfonylureas, DPP-4 inhibitors, GLP-1 receptor agonists, SGLT2 inhibitors, insulins, thiazolidinediones, alpha-glucosidase inhibitors, and meglitinides. For each class, an ever-exposure indicator was constructed as 1 if any drug exposure for that class was recorded prior to the baseline date, otherwise 0. The same matching code and class definitions were applied to both cohorts to ensure phenotype-equivalent medication features.

*Polygenic risk score selection.* We selected 20 polygenic risk scores from the PGS Catalog (<https://www.pgscatalog.org/>) covering ischemic stroke and clinically related cardiovascular and metabolic conditions. Scores were selected based on three criteria: (1) availability of computable weights in the catalog, (2) relevance to ischemic stroke risk either directly (stroke-specific scores) or through established stroke risk factors — hypertension, type 2 diabetes, lipid levels (LDL, HDL, total cholesterol, triglycerides), atrial fibrillation, coronary artery disease, and body mass index, and (3) variant coverage in both AoU whole-genome sequencing data and Bio*Me* TOPMed-imputed genotypes. Where multiple scores existed for the same trait, we selected the most recent and largest published score by training-set sample size. The primary ischemic stroke score used was PGS002724. The full list of selected scores with PGS Catalog identifiers, traits, training cohorts, and reference publications is in Supplementary Table 2.

*Race and ethnicity ascertainment.* In AoU, race and ethnicity were self-reported at enrollment via The Basics survey, using a single multi-select item ("Which categories describe you? Select all that apply") with response options: White; Black, African American, or African; Asian; American Indian or Alaska Native (AIAN); Middle Eastern or North African (MENA); Native Hawaiian or other Pacific Islander (NHPI); Hispanic, Latino, or Spanish; None of these fully describe me; and Prefer not to answer.

For modeling, each substantive response category was encoded as an independent binary indicator, preserving the multi-select structure rather than collapsing to a single category (e.g., a participant selecting both "White" and "Hispanic" had ethnicity_white = 1 and ethnicity_hispanic = 1). An additional binary indicator flagged participants selecting two or more substantive categories (ethnicity_multiracial). For participants without a Basics survey response, the Hispanic indicator was back-filled from the OMOP person.ethnicity_concept_id field ("Hispanic or Latino" = 1; "Not Hispanic or Latino" = 0; Skip / Prefer not to answer / None of these treated as missing).

For per-ancestry stratified analyses (Results, Figure 3, Supplementary Tables 10–12), single-category labels were derived hierarchically from these indicators: participants with ethnicity_hispanic = 1 were labeled Hispanic regardless of additional race selections; remaining participants with exactly one substantive race indicator were assigned that label (European American, African American, Asian, AIAN, MENA, NHPI); those with two or more non-Hispanic race indicators were labeled multiple/other.

In Bio*Me*, race and ethnicity were self-reported at recruitment using a comparable multi-select instrument and harmonized to the same binary-indicator and single-category coding conventions.

*Feature selection.* No manual feature selection or pre-filtering was applied beyond the structured feature engineering described above. XGBoost incorporates implicit feature selection through its tree-construction process — features that do not improve the loss-minimizing split at any node are effectively ignored — and we relied on this internal mechanism rather than imposing additional pre-filtering, which would risk discarding informative interactions that XGBoost can capture but univariate filters cannot.

*Tier composition.* The three nested model tiers used the following feature sets:

- **M1 (clinical baseline):** demographics, AoU surveys (or Bio*Me* equivalents), SNOMED CT-derived comorbidity indicators, and ever-exposure indicators across 29 cardiovascular and metabolic medication classes. M1 contains no biomarker measurements.
- **M2 = M1 + biomarker trajectories.** Adds longitudinal trajectory features (mean, standard deviation, slope, recency, ever-abnormal flag, fraction abnormal; defined above) derived from repeated measures of laboratory values, vital signs (systolic and diastolic blood pressure, heart rate), and anthropometrics (height, weight, body mass index).
- **M3 = M2 + polygenic risk scores.** Adds 20 PRS selected from the PGS Catalog as described above.

This tier structure enables clean separation of the marginal contributions of (i) repeated-measures biomarker information at the M2 tier and (ii) polygenic information at the M3 tier.

**Supplementary Methods 3. Definition of rFSRP-eligible and PCE-eligible subsets.**

Comparison of M3 with the Revised Framingham Stroke Risk Profile (rFSRP) and the Pooled Cohort Equations (PCE) required complete data on all variables specific to each score. Each traditional score was applied only to AoU test-partition participants meeting the relevant score's eligibility criteria and with complete data for all required inputs; missing values were not imputed for either traditional score.

*rFSRP-eligible subset (n = 1,338; 121 cases).* Eligibility required age ≥ 55 years and non-missing values for systolic blood pressure (most recent pre-baseline measurement), ever-exposure to antihypertensive medications, type 2 diabetes status, current cigarette smoking, atrial fibrillation history, and prior cardiovascular disease (defined as coronary artery disease, myocardial infarction, heart failure, or peripheral arterial disease). The most restrictive variable was the measured systolic blood pressure, which was available for 3,180 of 6,998 (45%) test-partition participants.

*PCE-eligible subset (n = 1,287; 83 cases).* Eligibility required age 40–79 years and non-missing values for self-reported race (Black vs non-Black, as PCE coefficients are not validated for other groups), total cholesterol (most recent pre-baseline value), HDL cholesterol (most recent), systolic blood pressure (most recent), ever-exposure to antihypertensive medications, type 2 diabetes status, and current cigarette smoking. Participants self-identifying as Black or African American were assigned the Black PCE coefficients; all other participants were assigned the non-Black PCE coefficients. The most restrictive variables were total cholesterol (2,038 / 6,998; 29% non-missing) and HDL cholesterol (1,761 / 6,998; 25% non-missing).

*Implications for interpretation.* Both subsets represent a more clinically active fraction of the test partition — participants with documented lipid panels and blood pressure measurements — which may bias the comparison cohort toward higher-risk individuals with greater EHR documentation. This selection effect does not invalidate the relative comparison between M3 and each traditional score within its eligible subset, but the absolute event rates and traditional-score AUROCs reported here should be interpreted accordingly.

*Note on PCE outcome alignment.* The Pooled Cohort Equations were originally derived to predict 10-year incidence of the atherosclerotic cardiovascular disease (ASCVD) composite outcome (nonfatal myocardial infarction, coronary heart disease death, and fatal or nonfatal stroke). We applied the published PCE coefficients to predict the stroke-only outcome used in our development cohort, which is expected to produce modestly lower discrimination than applying PCE to the full ASCVD composite. The reported PCE AUROC of 0.656 in our eligible subset is consistent with this expected attenuation and with reports of degraded PCE discrimination in contemporary multiethnic cohorts.

**Supplementary Methods 4. Discrimination, calibration, and threshold-dependent metric definitions.**

*Discrimination.* The area under the receiver operating characteristic curve (AUROC) summarizes a model's ability to rank cases higher than controls and is invariant to outcome prevalence; an AUROC of 0.5 indicates random ranking and 1.0 indicates perfect ranking. The area under the precision-recall curve (AUPRC) summarizes precision (positive predictive value) achieved across the full range of recall (sensitivity) and is more sensitive to class imbalance than AUROC; baseline AUPRC equals the outcome prevalence.

*Calibration.* The aggregate observed-to-expected (O/E) event ratio is the total number of observed events divided by the sum of predicted event probabilities; an O/E of 1.0 indicates perfect aggregate calibration, < 1.0 indicates net overprediction, and > 1.0 indicates net underprediction. The calibration intercept and slope are the intercept and slope estimates from a logistic regression of the observed binary outcome on the logit of the predicted probability. A calibration intercept of 0 and slope of 1 indicate perfect calibration. A nonzero intercept with slope near 1 corresponds to intercept-shift miscalibration — a uniform additive offset in log-odds space that preserves relative risk ordering but shifts the absolute level. This pattern is correctable by re-fitting the intercept on the target cohort without modifying the underlying model coefficients.

*Threshold-dependent metrics.* At the per-cohort 90%-specificity threshold, we report sensitivity (proportion of true cases correctly classified as high-risk), positive predictive value (PPV; proportion of high-risk classifications that are true cases), and negative predictive value (NPV; proportion of below-threshold classifications that are true controls). The 90%-specificity threshold was re-derived independently within each cohort to reflect cohort-specific event prevalence and predicted-probability distribution; it was not transferred between cohorts.

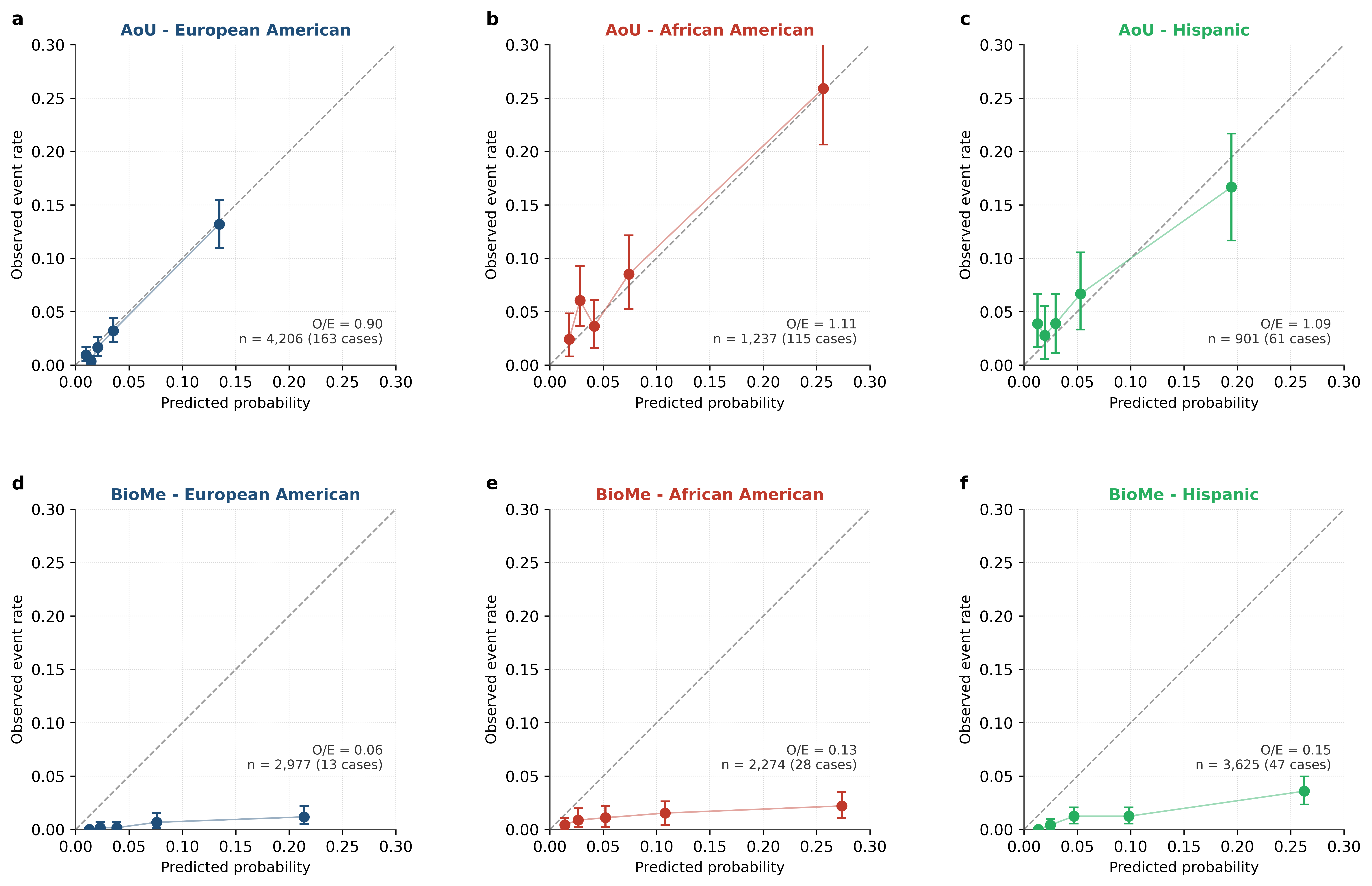

**Supplementary Figure 1. Calibration of the Full-cohort race-blind M3 model by ancestry stratum in the AoU test set (top row) and BioMe validation cohort (bottom row).**

For each cell, participants were divided into five equal-frequency bins by predicted 10-year ischemic stroke probability. Within each bin, mean predicted probability is shown on the x-axis and observed event rate on the y-axis, with vertical bars indicating 95% percentile-bootstrap confidence intervals (1,000 resamples). The dashed diagonal indicates perfect calibration (predicted = observed). The observed-to-expected (O/E) ratio reported in each panel summarizes aggregate calibration; a value of 1.0 indicates perfect aggregate calibration. AoU panels show points clustering along the diagonal across all three ancestries (O/E 0.90–1.11). BioMe panels show points sitting consistently below the diagonal across all three ancestries (O/E 0.06–0.15), indicating systematic overprediction of stroke risk in the BioMe cohort. The parallel-below-diagonal pattern is consistent with an intercept-shift miscalibration correctable by site-specific recalibration of the Platt scaling step. M3 = baseline clinical features + laboratory feature trajectories + polygenic risk scores; O/E = observed-to-expected event ratio.

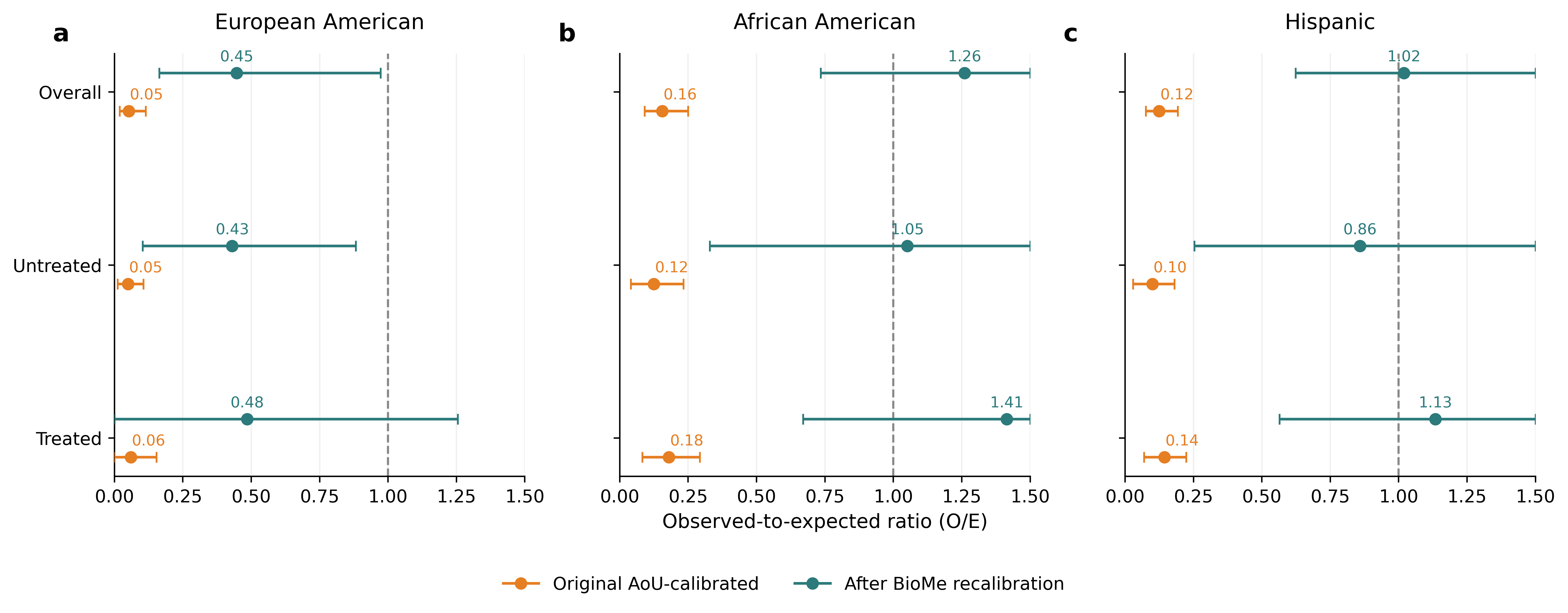

**Supplementary Figure 2.** **Treatment-stratified observed-to-expected ratios (O/E) of the harmonized full-cohort race-blind M3 model in the Bio*Me* validation cohort, before and after intercept-only recalibration.**

Each panel shows O/E values overall and stratified by baseline cardiovascular medication exposure (defined as any-ever use of statins, ACE inhibitors, angiotensin receptor blockers, antiplatelet agents, warfarin, factor Xa inhibitors, or direct thrombin inhibitors) for European American (a), African American (b), and Hispanic (c) participants. Orange markers show O/E from the original AoU-trained model with AoU-fit Platt calibration; teal markers show O/E after applying an intercept-only logistic recalibration fit on a stratified 50% partition of the Bio*Me* cohort and evaluated in the held-out 50%. Error bars are 95% percentile-bootstrap confidence intervals (2,000 resamples). The dashed vertical line at O/E = 1.0 indicates perfect aggregate calibration. The recalibration shifted O/E substantially toward unity in African American (overall O/E 0.16 → 1.26) and Hispanic (0.12 → 1.02) participants but produced incomplete correction in European American participants (0.05 → 0.45), indicating that per-ancestry recalibration may be more appropriate than a single global correction. Within each ancestry, treated and untreated participants showed similar O/E values, indicating that the cross-cohort calibration drift was not primarily explained by baseline cardiovascular medication exposure. M3 = clinical baseline + laboratory feature trajectories + polygenic risk scores.

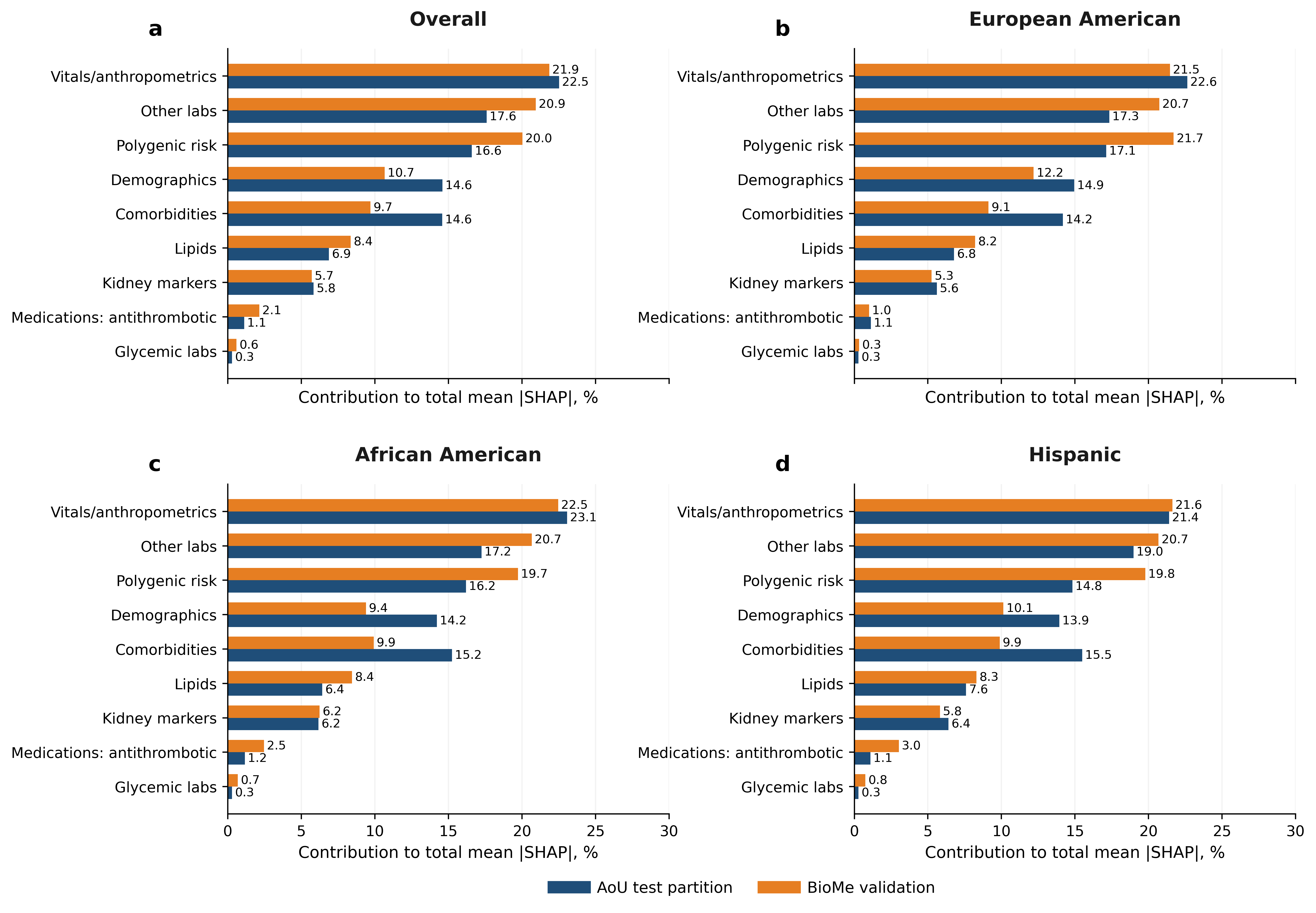

**Supplementary Figure 3.** **Domain-level Shapley additive explanation (SHAP) attribution of the harmonized full-cohort race-blind M3 model in the AoU test partition (blue) and Bio*Me* validation cohort (orange).**

For each model feature, mean absolute SHAP values were computed at the per-prediction level and aggregated within nine clinical feature domains. Values shown are the percentage contribution of each domain to total mean |SHAP|, ranked top-to-bottom by overall Bio*Me* contribution. Panel a shows the overall cohort; panels b–d show the same comparison restricted to participants self-identifying as European American (b), African American (c), and Hispanic (d). Polygenic risk score contributions were higher in Bio*Me* than AoU across all ancestry strata, with the largest cross-cohort difference observed in Hispanic participants (19.8% vs 14.8%). Comorbidity and demographic domain contributions were correspondingly lower in Bio*Me* than in AoU. Vitals and anthropometrics were the leading contributors in both cohorts (~22%). M3 = clinical baseline + laboratory feature trajectories + polygenic risk scores.
